# Machine learning approaches for fracture risk assessment: a comparative analysis of genomic and phenotypic data in 5,130 older men

**DOI:** 10.1101/2020.01.09.20016659

**Authors:** Qing Wu, Fatma Nasoz, Jongyun Jung, Bibek Bhattarai, Mira V Han

**Author notes:** **Correspondence:** Qing Wu, MD, ScD, Nevada Institute of Personalized Medicine, Department of Environmental and Occupational Health, School of Public Health, University of Nevada Las Vegas, 4505 Maryland Parkway, Las Vegas, NV 89154-4009.

## Abstract

The study aims were to develop fracture prediction models by using machine learning approaches and genomic data, as well as to identify the best modeling approach for fracture prediction. The genomic data of Osteoporotic Fractures in Men, cohort Study (*n* = 5,130), was analyzed. After a comprehensive genotype imputation, genetic risk score (GRS) was calculated from 1,103 associated SNPs for each participant. Data were normalized and split into a training set (80%) and a validation set (20%) for analysis. Random forest, gradient boosting, neural network, and logistic regression were used to develop prediction models for major osteoporotic fractures separately, with GRS, bone density and other risk factors as predictors. For model training, the synthetic minority over-sampling technique was used to account for low fracture rate, and 10-fold cross-validation was employed for hyperparameters optimization. In the testing set, the area under the ROC curve (AUC) and accuracy were used to assess the model performance. The McNemar test was employed for pairwise comparisons to examine the accuracy difference between models. The results showed that the prediction performance of gradient boosting was the best, with AUC of 0.71 and an accuracy of 0.88, and the GRS ranked as the 7th most important variable in the model. The performance of random forest and neural network were also better than that of logistic regression. Pairwise comparisons showed that the accuracy difference between models was significant. This study suggested that improving fracture prediction can be achieved by incorporating genetic profiling and by utilizing the gradient boosting approach.

## Introduction

Osteoporotic fractures continue to be a significant, ever-growing public health problem around the world. By estimation, more than 8.9 million fractures worldwide are caused by osteoporosis every year [1]. The incidence of osteoporotic fracture increases exponentially throughout life, as does the risk of devastating consequences of these fractures, including functional decline, institutionalization, mortality, and destitution. With longevity increasing globally, the potentially high cumulative rate of osteoporosis and fracture, and the associated excess disability and mortality [2], have caused and will continue to cause an inevitable increase in social and economic burdens worldwide.

Osteoporosis affects both men and women. Women have a higher risk of osteoporotic fracture, and thus the general population and even the medical community share the belief that osteoporosis is not as important in men. Therefore, osteoporosis has been an under-recognized problem, and as well, osteoporotic fracture has become a long-neglected medical issue in men. Men indeed suffer much higher morbidity and mortality rates than women following osteoporotic fractures [3]. For example, men are more likely to die from hip fractures, with a mortality rate in men of up to 37.5% [4]. Of those survivors, men are significantly more likely than women to have disabilities and lose independence. With increasing longevity in men, the burdens resulting from their fractures are likely to increase significantly shortly.

Therefore, accurately identifying high-risk individuals is critical to preventing fracture and the subsequent devastating consequences, especially in older men. Primary risk factors contributing to fracture susceptibility of men include advancing age, low bone mineral density (BMD), low body weight, limitations of physical function, previous fracture, a history of falls, prolonged use of corticosteroids, smoking, and alcohol consumption [5]. Several predictive models have developed based on these risk factors, with the Fracture Risk Assessment Tool (FRAX) being the most commonly used fracture prediction tool [6 - 7]. However, the prognostic accuracy of FRAX is suboptimal, because the average area under the receiver operating characteristic curve (AUC); the primary metric for model assessment for total fracture by FRAX is only 0.67 (95% CI, 0.64-0.71) [10]. Thus there is room for further improvement in fracture prediction.

Substantial efforts have begun to find ways to improve fracture prediction in the field. Generally, integrating new markers of fracture risk in the prediction model and adopting innovative modeling strategies are two essential approaches to improving the accuracy of fracture prediction. One crucial factor improving the accuracy of fracture risk assessment is genetic factors, which have not been included in existing fracture risk assessment models [7]. However, genetic elements are determinants of bone structure and predisposition to bone deterioration and fragility. Mounting evidence shows that fracture susceptibility is genetically determined [8 - 10]. In the last two decades, major genome-wide association studies (GWASs) have successfully identified thousands of single nucleotide polymorphisms (SNPs) associated with fracture [10 - 11]. To date, the largest GWAS on fracture identified 1,103 SNPs associated with fracture [10]. Integrating these SNPs into fracture risk assessment models has the potential to improve the accuracy of fracture prediction over the existing models.

Additionally, existing models for fracture prediction do not take into account the potential interactions between risk factors, which are likely present. Such limitations can be addressed by machine learning (ML) approaches, which is capable of modeling complex interactions and maximizing predictive accuracy from complex data. ML techniques, including random forest, gradient boosting, and neural network, have been applied in clinical research for disease predictions and have shown much higher accuracy for diagnosis than classical methods [12]. However, the performance of these ML techniques for fracture prediction has rarely been examined.

Integrating genomic data and using these ML techniques will likely enable the development of a novel model to improve fracture prediction. Therefore, the aims of the present work were 1) to develop models using ML techniques to predict major osteoporotic fracture (MOF) in men, based on data containing genomic variants; 2) To compare these models to determine which ML model performs the best for MOF prediction.

## Materials and Methods

### Data Source

The existing data from the Osteoporotic Fractures in Men Study (MrOS) archived in the Database of Genotypes and Phenotypes (dbGaP) was used for the analysis. Genotype and phenotype data of MrOS was acquired through authorized access (Accession: phs000373.v1.p1) after the analysis plan was approved by the institutional review board at the University of Nevada, Las Vegas, and the National Institute of Health (NIH). MrOS was designed to investigate anthropometric, lifestyle, and medical factors related to bone health in older, community-dwelling men. Details of the MrOS research design, recruitment, and baseline characteristics have been described elsewhere [13 - 14].

### Study participants

Participants in the MrOS were at least 65 years old, community-dwelling, ambulatory, and not having a bilateral hip replacement at the study entry. For enrollment, participants were required to be able to complete the self-administered questionnaire, to understand and sign the written informed consent, to attend the clinic visit, and to complete at least the anthropometric, DEXA, and vertebral X-ray procedures. The participants did not have a medical condition that would result in imminent death, a judgment that was made by the investigators. A total of 5,994 men were enrolled between March 2000 and April 2002; all were from six communities in the United States (Birmingham, AL; Minneapolis, MN; Palo Alto, CA; Pittsburgh, PA; Portland, OR; and San Diego, CA.). The selection process for the MrOS led to 5,130 eligible subjects, all of whom had both genotype data and phenotype data available for our current analysis.

### Measurements of BMD and quantitative ultrasound (QUS)

During the baseline visit of MrOS, total body, total femur, and lumbar spine (L1 to L4) BMD were measured using a fan-beam dual-energy X-ray absorptiometry (QDR 4500 W, Hologic, Inc., Bedford, MA, USA). Participants were scanned by centrally certified DXA Clinical Densitometrists, all of whom used standardized procedures for BMD measurements. Cross-calibrations found no linear differences across scanners, and the maximum percentage difference between scanners was only 1.4% in mean BMD of total spine [15]. No shifts or drifts in scanner performance were found, based on longitudinal quality evaluation from data that was daily scanned at each clinical center for standardized phantoms.

QUS measurements in Mr. OS were described in detail else [16]. Briefly, QUS was measured at the right heel for participants using the Sahara machine (Hologic, Waltham, MA). This device provides 3 QUS parameters: broadband ultrasonic attenuation (dB/MHz), speed of sound (SOS in M/sec), and quantitative ultrasonic index (QUI, a unitless proprietary linear combination of BUA and SOS). Based on duplicate measurements on participants, the mean coefficient of variation for all devices was only 3.3.

### Assessment of covariates

Self-administered questionnaires were used to acquire bone-health related information, including demographics, clinical history, medications, and lifestyle factors. The acquired information contained variables in the study, including age, race, smoking, and alcohol consumption. Height (cm) was measured using a Harpenden Stadiometer, and weight (kg) was measured by a standard balance beam or an electronic scale. BMI was calculated as kilograms per square meter. Smoking was categorized as “never,” “past,” and “current.” Alcohol intake was quantified in terms of the usual drinks consumed per day. Walking speed was determined by timed completion of a 6-meter course, traveled at the participant’s usual walking speed. Mobility limitations were measured by participants’ ability to rise from a chair without using their arms, as well as their ability to complete five chair stands.

### Major Osteoporotic Fracture

A major osteoporotic fracture (MOF) was defined as a fracture of the hip, spine (clinical), wrist, or humerus [17] that had occurred during the following up in this study. Only 451 men (8.8%) were found to have a MOF in the data.

### Genotyping Data

Baseline whole blood samples were used for DNA extraction. Written consent for DNA use was obtained in advance from the participants. Quality-control genotype data were acquired through dbGaP. A most comprehensive Haplotype Reference Consortium (HRC) reference panel and a Positional Burrows-Wheeler Transform (PBWT) imputing algorithm were utilized for the genotype imputation to ensure high quality of imputation. All genotype imputation work was conducted at the Sanger Imputation Service. Based on a most up-to-date study published by Morris et al. in 2019 [10], a total of 1,103 associated SNPs were extracted for this analysis. Each of the 1,103 SNPs were successfully imputed for MrOS data. The imputation quality was excellent, with a mean R^2^ of 0.99.

### Genetic risk score

A genetic risk score (GRS) is a standardized metric derived from the number of risk alleles and their effect size for each study participant. This metric allows the composite assessment of genetic risk in complex traits. A linkage disequilibrium (LD) pruning was performed in advance in order to eliminate possible LD between SNPs. None of the 1,103 SNPs was removed after the pruning. The weighted GRS was then calculated with the algorithms described else [18]. Briefly, each individual’s weighted GRS was calculated by summing the number of risk alleles at each locus and multiplied by the effect size, which provided regression coefficients from the referenced study [10].

### Data analysis

In the phenotype dataset, only the ultrasound speed of sound had missing values (6.7%). Median imputation, a most common imputation method for phenotype data, was used to replace these missing values so as to maximize the sample size. Phenotype data (*n* = 5,130) and genotype data (*n* = 5,143) were merged after all missing data imputations were completed. The predictors included GRS, age, race, body weight, height, smoking, alcohol consumption, walking speed, impairment of instrumental activities of daily living, BMD, mobility limitations, and ultrasound speed of sound. All modeling was adjusted for clinical location. We included the three BMD measurements from the femoral neck, total spine, and total hip in order to predict MOF in the models. Since site-specific measures of BMD had the strongest association with fracture at the corresponding site [20 - 23], including multiple BMD measurements should have been a better prediction for MOF, which includes fractures from multiple skeletal sites. Our analysis also confirmed that multiple BMD measurements had better MOF predictions than single BMD in the model. Centered predictors were used to address possible multicollinearity problems between multiple BMD predictors in the models. All continuous variables in the data were normalized by the centering method before analysis. Centering independent variables removes nonessential ill-conditioning and thus reduces multicollinearity. In this analysis, all variance inflation factors (VIF), which measured the impact of collinearity among the variables in the model, were < 5 after centering in logistic regression (VIF < 10 is considered acceptable). Advanced ML methods are more robust to multicollinearity [23]. Among the three very highly correlated QUS measurements [16], we only included SOS in the models. Because after centering, our analysis confirmed that SOS had the best MOF prediction performance, and including all three QUS measurements in the model did not improve the model performance. The final dataset (*n* = 5,130) was divided randomly into the training set (80%, *n* = 4,104) and the testing set (*n* = 1,026). The data processing flow and corresponding sample size in each step is shown in Figure 1.

**Figure 1.**
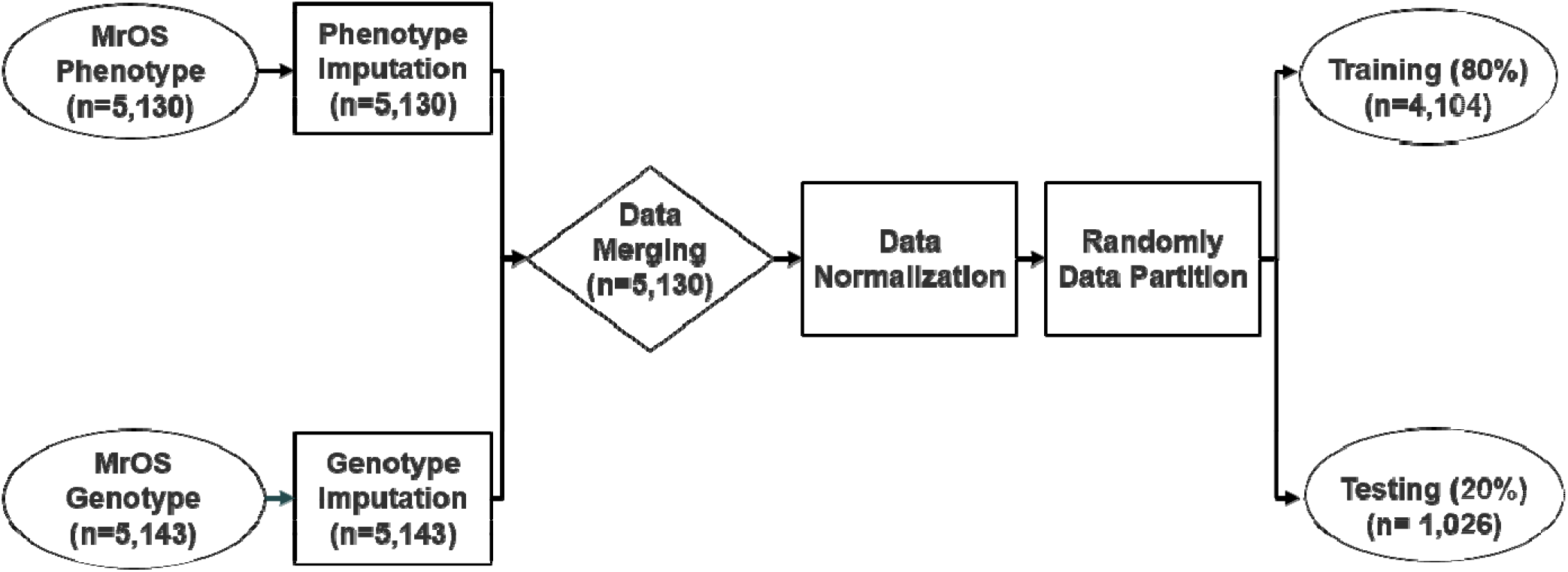
Overview of Data Process Flow

The MrOS had a small number of fracture cases (*n* = 451, 8.8%). Such imbalanced data could cause difficulty in using accuracy of prediction to assess model performance because the minor class (fracture) should have less influence on accuracy than the major class (non-fracture) [24], which could contribute to the undesirable lower performance of the prediction model [25]. To address this data imbalance issue, we employed the Synthetic Minority Over-sampling (SMOTE) technique [26] for data resampling in order to account for the low fracture rate in the training set. We used 10-fold cross-validation for hyper-parameters optimization, for which the training set (*n* = 4,104) is divided into 10-folds, with one fold chosen for validation and remaining folds used for training. We used Scikit-learn’s randomized search cross-validation method to find the best hyperparameters for various ML algorithms. Fracture prediction models were trained separately using logistic regression, random forest, gradient boosting, and neural network with backpropagation in the training set.

### Model evaluation

The testing set (*n* = 1,026) was used for model evaluation and comparison. The selected evaluation metrics included the area under the ROC curve (AUC) and the prediction accuracy. The receiver-operating characteristic (ROC) curve is generated by plotting true positive rate vs. false positive rate at various threshold settings. The prediction accuracy is the rate of correct predictions by each model. We employed the McNemar statistical test for pairwise comparisons for prediction accuracy between ML models [27]. We also used the testing set to investigate the variable importance in the best prediction model identified in this study. All of the analyses were performed in the Python Software Foundation, Python Language Reference (v3.7.3) with the package Scikit-learn: Machine Learning in Python (http://www.python.org) [28] was used.

## Results

### Baseline characteristics

Table 1 shows the characteristics of participants with MOF (*n* = 451) and without MOF (*n* = 4,679). The mean age of participants with MOF was significantly higher (75.9 years) than those without MOF (73.6 years). Each BMD measured from the femoral neck, total hip, and total spine in participants with MOF was significantly lower than that of the participants without MOF. The ultrasound speed of sound was lower in participants with MOF than that in participants without MOF, although the difference is not statistically significant.

**Table 1.** Demographic and Clinical Characteristics of participants with major osteoporotic fracture (MOF) in the follow up in the MrOS study.

| **Variable\*** | **With MOF**<br>**(n = 451)** | **Without MOF**<br>**(n = 4,679)** | **P-value\*\*** |
| --- | --- | --- | --- |
| **Age** | 75.9 ± 6.2 | 73.6 ± 5.8 | < 0.001 |
| **Weight** | 80.6 ± 13.1 | 83.4 ± 13.3 | < 0.001 |
| **Height** | 173.7 ± 6.6 | 174.2 ± 6.8 | < 0.001 |
| **Alcohol Consumption** | 3.9 ± 5.7 | 4.2 ± 6.8 | < 0.001 |
| **Impairment of**<br>**Instrumental**<br>**Activities of Daily**<br>**Living** | 0.5 ± 1.0 | 0.4 ± 0.9 | < 0.001 |
| **Femoral Neck BMD** | 0.7 ± 0.1 | 0.8 ± 0.1 | < 0.001 |
| **Total Hip BMD** | 0.9 ± 0.1 | 1.0 ± 0.1 | < 0.001 |
| **Total Spine BMD** | 1.0 ± 0.2 | 1.1 ± 0.2 | < 0.001 |
| **Ultrasound Speed of**<br>**Sound** | 1539.4 ± 31.8 | 1556.5 ± 35.8 | 0.14 |
| **Generic Risk Score** | 31.6 ± 0.4 | 31.6 ± 0.5 | < 0.001 |
| **Walking speed** | 1.0 ± 0.3 | 1.1 ± 0.3 | < 0.001 |
| **Mobility Limitations** | 0.3 ± 0.6 | 0.2 ± 0.5 | < 0.001 |
| **Race (White)** | 404 (90%) | 4212 (91%) | < 0.001 |
| **Smoking (Current)** | 275 (61) | 2742 (91) | < 0.001 |

\* Continuous variables were expressed as mean ± SD, and categorical variables were expressed as number (%).
\*\* *p* - values were obtained by *t*- test for continuous variables and chi-square test for categorical variables.

### Model Performance Comparison

Random forest, gradient boosting, neural network, and logistic regression models were developed with 10-folds cross-validation in the training set (*n* = 4,104), and their fracture prediction performance was compared in the testing set (*n* = 1,026). Figure 2 shows the comparison of the ROC curve of these developed models in the testing dataset for MOF prediction. The ROC curves of random forest, gradient boosting, and neural network are above that of the logistic regression. Figure 3 shows the performance results of each model in predicting MOF. AUC and accuracy of random forest, gradient boosting, and neural network are higher than those in the logistic regression model.

**Figure 2.**
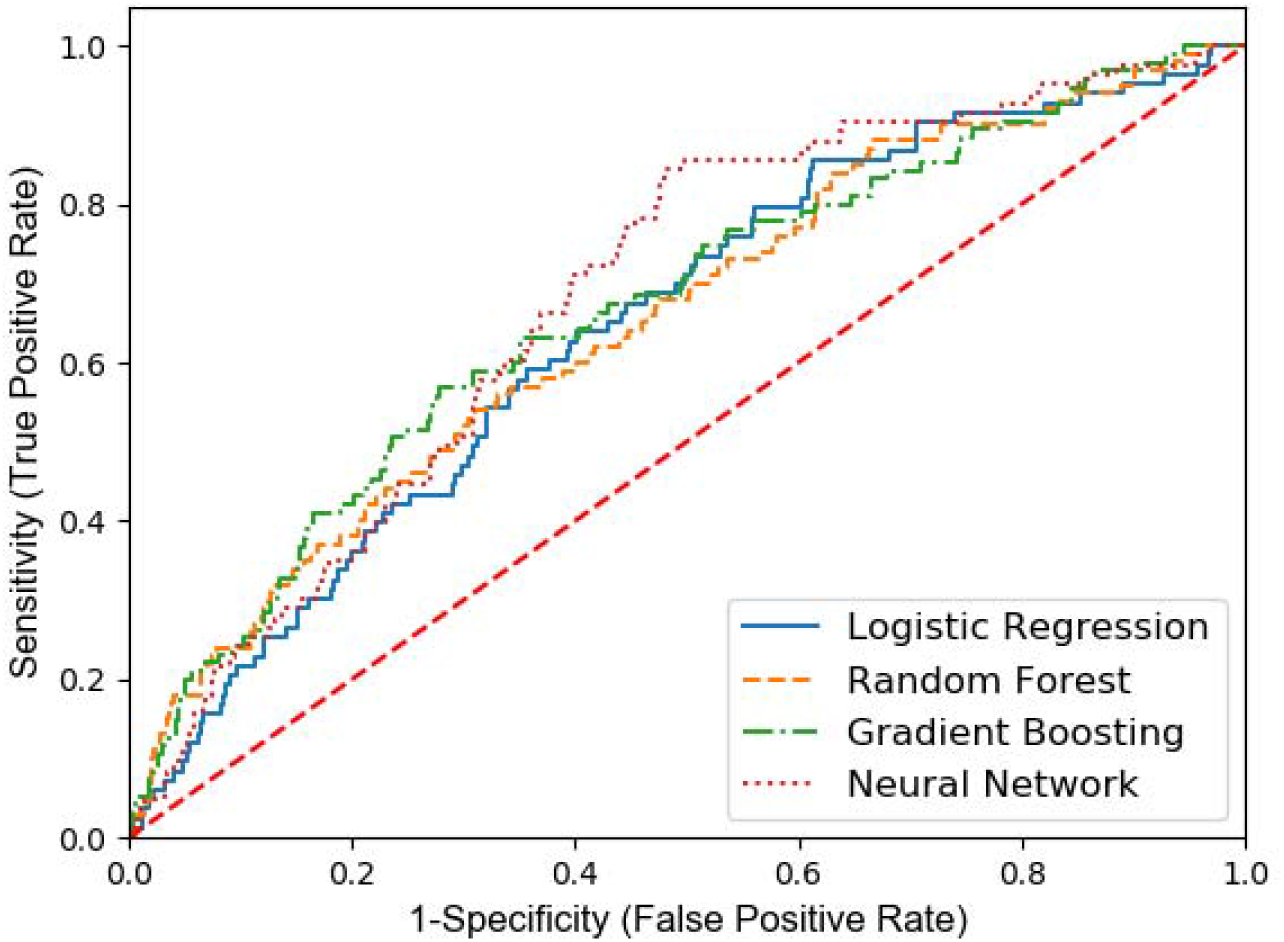
Comparisons of Receiver-operating characteristic curve between various ML models for MOF prediction in the testing dataset (n = 1,026)

**Figure 3.**
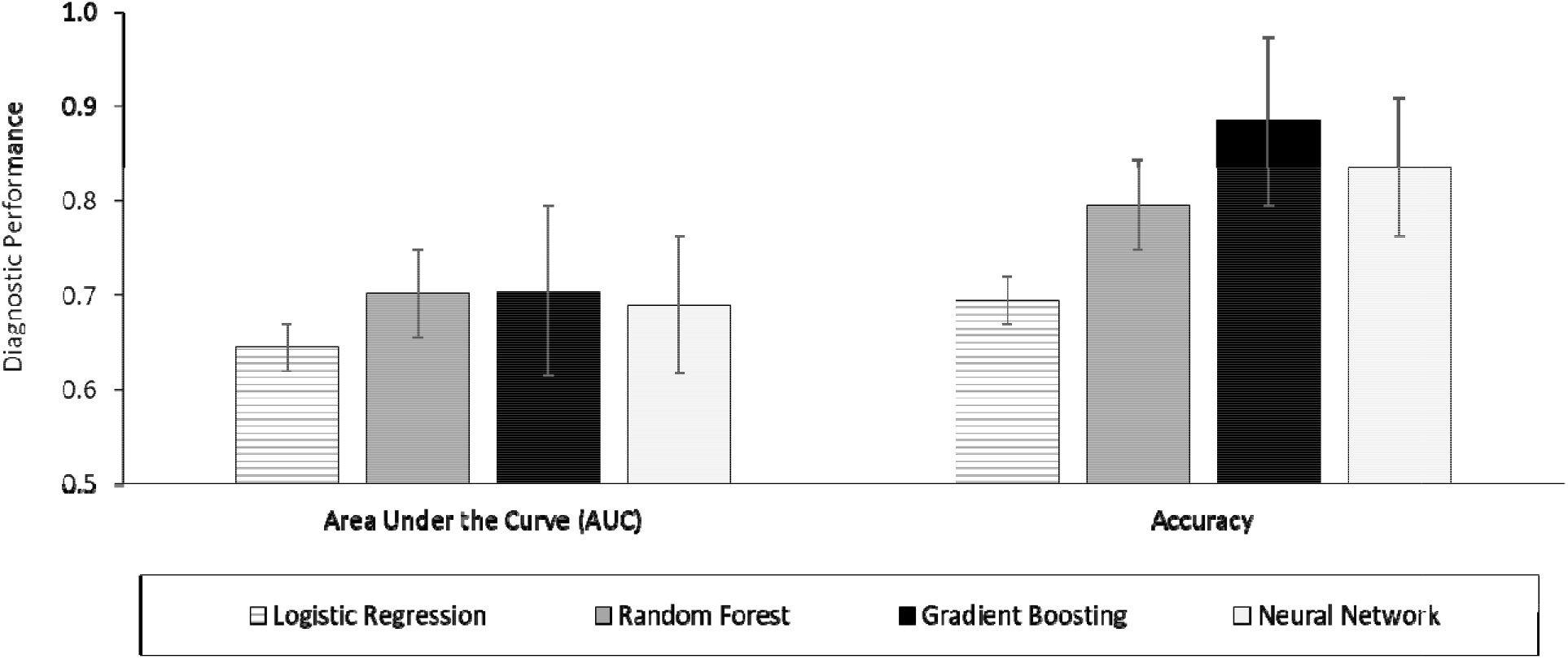
Diagnostic performance between various ML models in predicting Major Osteoporotic Fracture (MOF) in the testing dataset (n = 1,026)

For comparing each pair of models (e.g., Random Forest vs. Logistic Regression), we calculated the counts that made correct and incorrect predictions by each model. Table 2 shows the results of the McNemar test for comparison accuracies in classifying MOF (yes vs. no) between ML methods in the testing set. With bonferroni correction for multiple comparisons (α = 0.05/6=0.0083), the difference of accuracy between two models in all pairwise comparisons was statistically significant with *p* < .0001, except the comparison between logistic regression vs. neural network, with *p* = .025.

**Table 2.** Results of the McNemar’s Test between various ML models in predicting Major Osteoporotic Fracture in the testing dataset (n = 1,026)

| **Cross-Validation**<br>**Model** | ***p* Value** |  |  |
| --- | --- | --- | --- |
|  | **Logistic**<br>**Regression** | **Random**<br>**Forest** | **Gradient**<br>**Boosting** |
| **Neural**<br>**Network** | < .05 | < .0001 | < .0001 |
| **Gradient**<br>**Boosting** | < .0001 | < .0001 | -- |
| **Random**<br>**Forest** | < .0001 | -- | -- |

### Assessment of Variable Importance

Using the testing set, we investigated the variable importance in the gradient boosting model, which was the optimal prediction model identified in this study. Figure 4 shows the mean decrease in the Gini impurity of gradient boosting. Total hip BMD, femoral neck BMD, SOS, Age, total spine BMD, Weight, GRS, and height were the most important variables for the MOF prediction in the gradient boosting model. The GRS is ranked as the 7*^th^* most important variable in the model.

**Figure 4.**
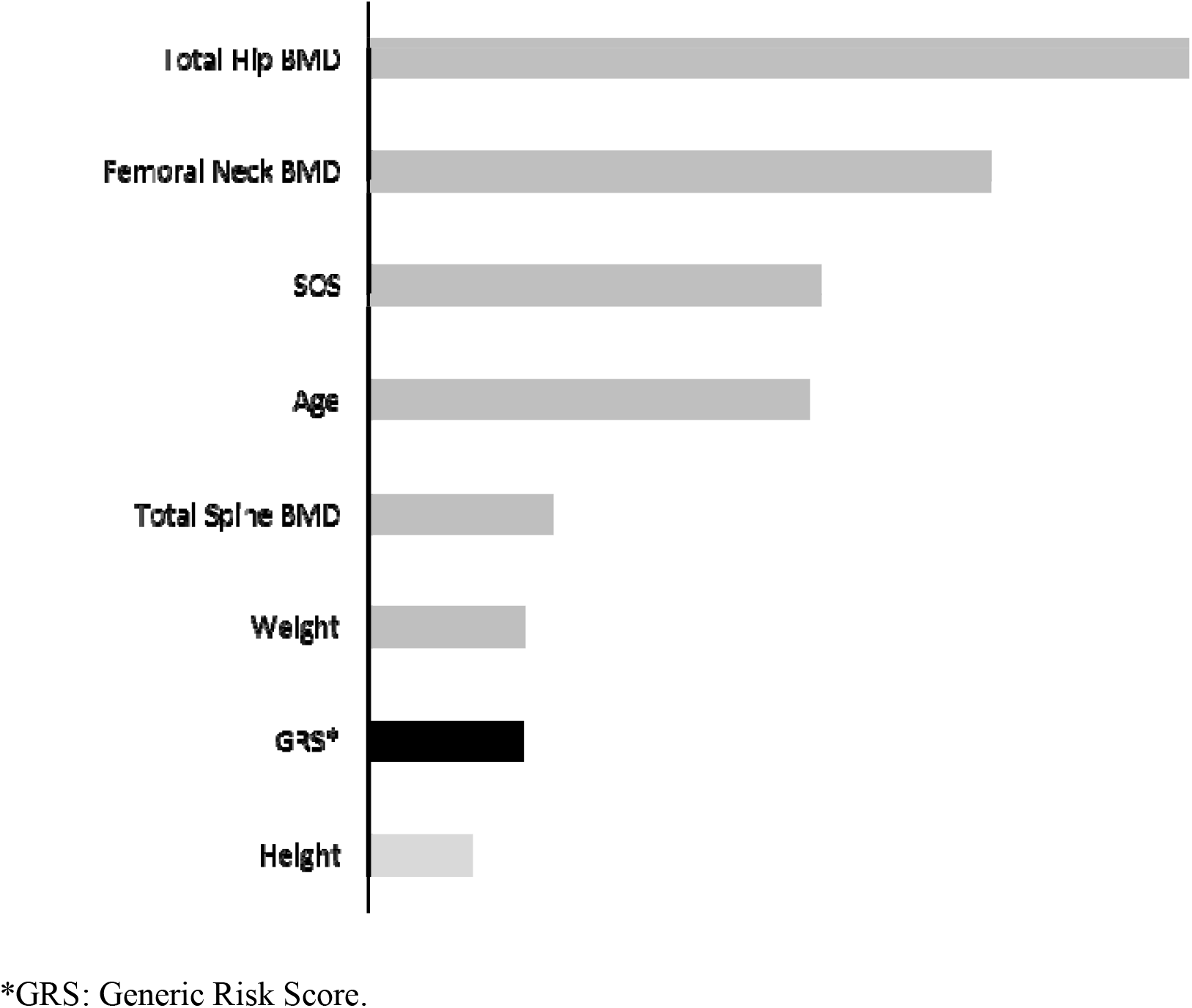
Variable importance in the gradient boosting model for prediction of Major Osteoporotic Fracture in the testing set (n = 1,026).

## Discussion

This study presents the finding that using various ML approaches to develop the predictive model for MOF by analyzing both genotype and phenotype data. The study results demonstrate that several ML approaches perform better than logistic regression and that the gradient boosting model appears to be the best to predict MOF in men. The present study also demonstrates that GRS is an important variable for fracture prediction and that GRS provides information on osteoporotic fracture risk, which is complementary to BMD and other risk factors. As existing models for fracture risk assessment are suboptimal, for example, FRAX, which is widely accepted worldwide, only have AUC value around 0.7 for total fracture prediction [29]. The prediction performance of other developed models for fracture risk assessment, including Garvan Fracture Risk Calculator [29], is not better. There is a crucial need to find ways to improve prediction accuracy and performance for fracture risk. The present study demonstrates that fracture prediction accuracy can be improved by incorporating genetic profiling and utilizing advanced ML modeling approaches. To the best of our knowledge, our present work is the first attempt to predict fracture outcomes using both advanced ML approaches and genetic information, as well as to identify the best performing model for MOF prediction. As such, this study will do much to facilitate finding a new, personalized tool to assess individual fracture risk.

Genetic factors that influence osteoporotic fracture risk is well documented. Hereditary factors contribute 50%-85% to fracture susceptibility [30]. Several large GWAS and GWAS meta-analyses have already identified more than 1,000 SNPs associated with fracture risk at genome-wide significant levels [10 - 11]. Although these individual SNPs have modest effect size on fracture risk, GRS, as summarized from individual risk SNPs, can improve AUC and accuracy of fracture prediction [31]. In the present study, we found that GRS ranked the 7^th^ most important variable in the optimal MOF prediction model of gradient boosting, where the model includes major conventional risk factors as predictors. The contributions of GRS to fracture prediction we observed in this study were consistent with previous studies [31], which used 63 associated SNPs identified 8 years ago [32]. We utilized the most updated 1,103 recently discovered associated SNPs [10] for this analysis. Our study further supports that genetic profiling has great potential for improving the accuracy of fracture prediction over and above the existing models, which only employ conventional phenotypic risk factors.

Of equal importance, advanced ML approaches provide great potential for improving fracture prediction. The reason is that besides disregarding genetic factors, most, if not all, existing fracture prediction models do not allow for potential interactions between predictors. However, interactions between predictors are likely present but not detected by conventional modeling approaches. Such weakness in existing models can be remedied with the advanced ML techniques we are exploring in our current research. In the present study, we employed multiple ML approaches, including random forest, gradient boosting, and neural networks, in developing a more accurate fracture prediction model. We found that gradient boosting has the highest accuracy and AUC for MOF prediction in our testing set, and the accuracy of the gradient boosting model is significantly higher than that of the other three models, as verified by the McNemar test, a widely used approach for comparing ML classifiers. The highest predictive performance of the gradient boosting model has been widely used in other areas for various outcomes, including urinary tract infections [33], hip fractures [34], hepatocellular carcinoma [35], sepsis [36], and bioactive molecules [37]. The present study suggested that the gradient boosting algorithm, combined with genomic profiling as a predictor, can provide a more accurate prediction for fracture risk.

Our study has limitations. First, the total sample size (*n* = 5,130) is relatively small for ML approaches. ML approaches often require much larger training data size. For this reason, we employed a 10-folds cross-validation approach for turning the hyper-parameter within the training set, instead of allocating part of the study sample for validation purpose, which would cause a smaller sample size for ML model training. Second, some covariates were not available in the MrOS data through dbGaP, including medications, comorbidities, and physical activities. A lack of these predictor variables can decrease the model performance. Studies with larger sample sizes may have found that the gradient boosting model had better prediction performance, with higher AUC [19, 25] than that observed in this study. Lack of related predictors in the data may explain lower AUC in the present study. The limited number of fracture cases is another reason for the lower AUC observed in this study. A testing dataset with at least 75 – 100 cases are recommended in order to achieve the consistent precision performance for an ML algorithm[38]; however, our testing dataset only had only 60 fracture cases. Third, the MrOS data we used only included men ≥ 65 years, so our findings may not apply to women or to individuals who are of a younger age. Fourth, rare risk SNPs were less likely to be included in this study, because risk SNPs used in this study were identified from a GWAS study, which likely discovered common variants, not rare variants [39]. Finally, due to the small sample size and a small number of hip fracture cases (*n* = 188) in MrOS data, we were not able to develop a predictive model for hip fracture outcomes by utilizing oversampling techniques. For a similar cause, we were not able to model 1,103 individual SNPs during the ML model development or to examine the interactions between these SNPs. We have to use GRS of these risk SNPs in the model due to the small number of fracture cases and the small sample size of this study. Nevertheless, these limitations are unlikely to have altered our findings in the present study because this is a self-control study, with all ML models developed and validated by the same datasets.

In summary, advanced ML models performed better in fracture prediction for men than conventional logistic regression. Gradient boosting appears to be the best performing model for MOF prediction in this study. As well, genetic variants contribute to the fracture prediction independent of BMD and other risk factors. Hence, our work suggests that improving fracture prediction accuracy can be achieved by incorporating genetic profiling and by utilizing the gradient boosting ML approach. Additional more extensive and more comprehensive studies, especially those including women, young participants (<65 years), rare genetic variants, and additional risk factors, are warranted in order to further examine fracture prediction performance of ML models, especially with individual SNPs as predictors. Genetically-enhanced, highly accurate assessment models are likely to improve fracture prediction and thus help clinicians and patients to assess fracture risk better at the individual level.

## Data Availability

The existing data from the Osteoporotic Fractures in Men Study (MrOS) archived in the Database of Genotypes and Phenotypes (dbGaP) was used for the analysis. Genotype and phenotype data of MrOS was acquired through authorized access (Accession: phs000373.v1.p1) after the analysis plan was approved by the institutional review board at the University of Nevada, Las Vegas, and the National Institute of Health (NIH).

**Abbreviations**
MrOS: Osteoporotic Fractures in Men Study
ML: Machine Learning
BMD: Bone Mineral Density
FRAX: The Fracture Risk Assessment Tool
GRS: Generic Risk Score
QUS: Quantitative Ultrasound
ROC: Receiver Operating Curve
AUC: Area Under Curve
LR: Logistic Regression
RF: Random Forest
GB: Gradient Boosting
NN: Neural Network
MOF: Major Osteoporotic Fracture
SNPs: Single Nucleotide Polymorphisms
FNBMD: Femoral Neck BMD
TSBMD: Total Spine BMD
THBMD: Total Hip BMD

## Acknowledgments

The data/analyses presented in the current publication are based on the use of study data downloaded from the dbGaP web site, under phs000373.v1.p1 (https://www.ncbi.nlm.nih.gov/projects/gap/cgi-bin/study.cgi?study_id=phs000373.v1.p1). The research and analysis described in the present study was supported by a COBRE grant from the National Institute of General Medical Sciences (GR08954), the Genome Acquisition to Analytics (GAA) Research Core of the Personalized Medicine Center of Biomedical Research Excellence at the Nevada Institute of Personalized Medicine, and the National Supercomputing Institute at the University of Nevada Las Vegas. The funding sponsors were not involved in the analysis design, genotype imputation, data analysis, and interpretation of the analysis results or the preparation, review, or approval of this manuscript.

## References

[1] Johnell O, Kanis JA (2006) An estimate of the worldwide prevalence and disability associated with osteoporotic fractures. Osteoporosis International 17(12): 1726–1733

[2] Melton LJ, Cooper C (2007) Chapter 21 - Magnitude and Impact of Osteoporosis and Fractures, in Osteoporosis, 2nd ed., Academic Press Inc., 2007, 557–567

[3] Boonen S et al. (2012) Fracture Risk and Zoledronic Acid Therapy in Men with Osteoporosis. New England Journal of Medicine 367(18): 1714–1723

[4] Jiang HX et al. (2005) Development and initial validation of a risk score for predicting in-hospital and 1-year mortality in patients with hip fractures. Journal of Bone and Mineral Research 20(3):494–500

[5] Papaioannou A et al. (2009) Risk factors for low BMD in healthy men age 50 years or older: A systematic review. Osteoporosis International 20(4):507–518

[6] Kanis JA, Johnell O, Oden A, Johansson H, McCloskey E (2008) FRAX^TM^ and the assessment of fracture probability in men and women from the UK. Osteoporosis International 19: 385–397

[7] McCloskey E V., Johansson H, Oden A, Kanis JA (2009) From relative risk to absolute fracture risk calculation: The FRAX algorithm. Current Osteoporosis Reports 7(3):77–83

[8] Ralston SH, Uitterlinden AG (2010) Genetics of Osteoporosis. Endocrine Reviews 31(5):629–662

[9] Hsu YH et al. (2010) An integration of genome-wide association study and gene expression profiling to prioritize the discovery of novel susceptibility loci for osteoporosis-related traits. PLoS Genetics 6(6):1–16

[10] Morris JA et al. (2019) An atlas of genetic influences on osteoporosis in humans and mice. Nature Genetics 51(2):258–266

[11] Kim SK (2018) Identification of 613 new loci associated with heel bone mineral density and a polygenic risk score for bone mineral density, osteoporosis and fracture. PLoS ONE 13(7):e0200785

[12] Hsieh CH, Lu RH, Lee NH, Chiu WT, Hsu MH, Li YC (2011) Novel solutions for an old disease: Diagnosis of acute appendicitis with random forest, support vector machines, and artificial neural networks. Surgery 149(1):87–93

[13] Orwoll E et al. (2005) Design and baseline characteristics of the osteoporotic fractures in men (MrOS) study - A large observational study of the determinants of fracture in older men. Contemporary Clinical Trials 26: 569–585

[14] Blank JB et al. (2005) Overview of recruitment for the osteoporotic fractures in men study (MrOS). Contemporary Clinical Trials 26(5):557–568

[15] Cauley JA et al. (2005) Factors associated with the lumbar spine and proximal femur bone mineral density in older men. Osteoporosis International 16(12): 1525–1537

[16] Bauer DC, Ewing SK, Cauley JA, Ensrud KE, Cummings SR, Orwoll ES (2007) Quantitative ultrasound predicts hip and non-spine fracture in men: The MrOS study. Osteoporosis International 18(6):771–777

[17] Lix LM, Leslie WD, Majumdar SR (2018) Measuring improvement in fracture risk prediction for a new risk factor: A simulation. BMC Research Notes 1162

[18] Andrews NA (2010) Genome-wide association studies in the osteoporosis field: Impressive technological achievements, but an uncertain future in the clinical setting. IBMS BoneKEy 7(11):382–387

[19] Cummings SR et al. (1993) Bone density at various sites for prediction of hip fractures. The Lancet 341(8837):72–75

[20] Melton LJ, Atkinson EJ, O’Fallon WM, Wahner HW, Riggs BL (1993) Long-term fracture prediction by bone mineral assessed at different skeletal sites. Journal of Bone and Mineral Research 8(10):1227–1233

[21] Kanis JA et al. (2005) Assessment of fracture risk. Osteoporosis international: a journal established as result of cooperation between the European Foundation for Osteoporosis and the National Osteoporosis Foundation of the USA 16(6):581–9

[22] Katie L Stone, Dana G Seeley, Li-yung Lui, Jane A Cauley, Kristine Ensrud, Warren S Browner, Michael C Nevitt, Steven R Cummings F the study of osteoporotic fractures research group (2003) BMD at Multiple Sites and Risk of Fracture of Multiple Types: Long-Term Results From the Study of Osteoporotic Fractures Multiple Sites and Risk of Fracture of Multiple Types: Long-Term Results From the Study of Osteoporotic Fractures. Journal of Bone and Mineral Research 18(9): 1947–1954

[23] Iniesta R, Stahl D, McGuffin P (2016) Machine learning, statistical learning and the future of biological research in psychiatry. Psychological Medicine 46(12):2455–2465

[24] Sun Y, Kamel MS, Wong AKC, Wang Y (2007) Cost-sensitive boosting for classification of imbalanced data. Pattern Recognition 40(12):3358–3378

[25] Kotsiantis S, Kanellopoulos D, Pintelas P (2006) Handling imbalanced datasets: A review. GESTS International Transactions on Computer Science and Engineering 30(1):25–36

[26] Nitesh V. Chawla, Kevin W. Bowyer, Lawrence O. Hall WPK (2002) SMOTE: Synthetic Minority Over-sampling Technique Nitesh. Journal of Artificial Intelligence Research 16(1):321–357

[27] Raschka S (2018) Model Evaluation, Model Selection, and Algorithm Selection in Machine Learning. CoRR abs/1811.12808.

[28] Fabian Pedregosa, Gael Varoquaux, Alexandre Gramfort, Vincent Michel, Bertran Thirion, Olivier Grisel, Mathieu Blondel, Peter Prettenhofer, Ron Weiss, Vincent Dubourg, Jake Vanderplas, Alexandre Passos DC (2011) Scikit-learn: machine learning in Python. Journal of Machine Learning Research 122825–2830

[29] Bolland MJ et al. (2011) Evaluation of the FRAX and Garvan fracture risk calculators in older women. Journal of Bone and Mineral Research 26(2):420–427

[30] Eriksson J et al. (2015) Limited clinical utility of a genetic risk score for the prediction of fracture risk in elderly subjects. Journal of Bone and Mineral Research 30(1): 184–194

[31] Ho-Le TP, Center JR, Eisman JA, Nguyen HT, Nguyen T V. (2017) Prediction of Bone Mineral Density and Fragility Fracture by Genetic Profiling. Journal of Bone and Mineral Research 32(2):285–293

[32] Estrada K et al. (2012) Genome-wide meta-analysis identifies 56 bone mineral density loci and reveals 14 loci associated with risk of fracture. Nature genetics 44(5):491–501

[33] Taylor RA, Moore CL, Cheung KH, Brandt C (2018) Predicting urinary tract infections in the emergency department with machine learning. PLoS ONE 13(3): 1–15

[34] Kruse C, Eiken P, Vestergaard P (2017) Machine Learning Principles Can Improve Hip Fracture Prediction. Calcified Tissue International 100(4):348–360

[35] Sato M et al. (2019) Machine-learning Approach for the Development of a Novel Predictive Model for the Diagnosis of Hepatocellular Carcinoma. Scientific Reports 9(1):1–7

[36] Chiew CJ, Liu N, Tagami T, Wong TH, Koh ZX, Ong MEH (2019) Heart rate variability based machine learning models for risk prediction of suspected sepsis patients in the emergency department. Medicine 98(6):e14197

[37] Babajide Mustapha I, Saeed F (2016) Bioactive Molecule Prediction Using Extreme Gradient Boosting. Molecules (Basel, Switzerland) 21(8):1–11

[38] Beleites C, Neugebauer U, Bocklitz T, Krafft C, Popp J (2013) Sample size planning for classification models. Analytica Chimica Acta 760: 25–33

[39] Nguyen T V., Eisman JA (2013) Genetic profiling and individualized assessment of fracture risk. Nature Reviews Endocrinology 9(3): 153–161

